# Aerosols created by dental procedures in a primary care setting

**DOI:** 10.1101/2020.08.27.20183087

**Authors:** R S Hobson, S B Pabary, K Alamani, K Badminton

## Abstract

The study was undertaken to record the amount of dental aerosol created using 3-in-1 syringe, air rotor, and ultrasonic scaler using high volume suction (HVS) in 5 primary care dental surgeries. The time for the aerosol to dissipate following completion of the procedure was also recorded. The amount of aerosol created above the background level for the surgery corresponding to the operating positions of the nurse, dentist, and patient was recorded using particle meters measuring the number of 2.5µm (PM2.5) and 10µm (PM10) particles respectively. The procedures were recorded in triplicate for each surgery and average change calculated for each procedure, lasting 90 seconds.

PM2.5 remained at or very near background readings during all procedures, whereas PM10 increased with use of the air rotor and to a much lower extent with both 3-in-1, and ultrasonic scaler. The means time to return to background reading level was 2.5 minutes.

It was concluded that PM2.5 levels did not rise and although PM10 increased for all procedures the increase was low and with a return to background readings within 2m:50s (95% CI: 2:34 to 3:37) of completing the procedures that a minimum fallow period of 5minutes would allow be more than ample to be safe.

## Introduction

Aerosol has since the start of the COVID19 pandemic become a matter of concern and debate which is particularly important to the practice of dentistry (Meng et al., 2020, Peng et al., 2020, Su, 2020). Dental procedures routinely create a water / air mix or aerosol. But the quantity of aerosol and its potential role in respiratory borne viral disease transmission is unknown as the vast majority of previous studies into dental surgery contamination has been on blood, saliva, bacterial, and fungal splatter and surface contamination (Micik et al., 1969, Abel et al., 1971, Yamada et al., 2011, Polednik, 2014, Dawson et al., 2016, Kobza et al., 2018).

The concern is the potential transmission of virus during aerosol generating procedures (AGP). In medicine AGPs have been demonstrated as capable of respiratory virus transmission(Morgenstern, 2020). However, medical aerosols are fundamentally different from dental aerosols. A Medical Aerosol Generating Procedure (m-AGP) consists of 100% mucus, nasal and oral secretions with a high potential of carrying virus. A Dental Aerosol Generating Procedure (d-AGP) is the use of an instrument (water cooled handpiece, ultrasonic scaler, 3-in-1 water-air syringe) that introduces clean water into the oral cavity and hence becomes contaminated. The level of contamination of a d-AGP can be calculated to contain 0.04% of oral fluids (assuming homogenous mixing of saliva and water) it can therefore be assumed that the potential risk is significantly less than that of m-AGP. Current policies in dental practices in many countries have appear to have adopted the Precautionary Principle, where by if there non-zero probability of virus transfer at any level it has to be managed, even if there is no evidence to support the decisions (Taleb et al., 2020, WHO, 2020).

The level of potential exposure to air borne virus in the dental setting is driving the agenda for clinical procedures, cross infection control, and as the amount of aerosol created by d-AGPs in general dental practice remains poorly reported. This study was undertaken in a primary care setting to provide data relevant to the majority of dental practice.

## Materials and Methods

The study was undertaken in 3 dental practices in the North East of England recording the procedures in a number of surgeries being operated in their routine pre Covid-19 protocol.

Measurement of airborne aerosol particles is difficult and surrogates were use in the form of measuring PM2.5 (2.5 µm particles) as the smaller aerosol size associated with those more likely to remain airborne the longest and to be inhaled deeply to the respiratory tract. PM10 (10µm particles) as the larger aerosol that remain airborne for a shorter time period, more likely to merge and drop by gravity to contaminate surfaces, leading to fomite transmission of viral disease. PM2.5 is thought to be particle sizes most like to be associated with air borne transmission

One of the authors acted as the patient (SP, AK or KB), whilst a second author (RSH) operated the d-AGP instrument and high-volume suction. Each d-AGP instrument (3-in-1, air rotor and ultrasonic) was run for 90 seconds whilst a high-volume suction, held next to the lower left first molar throughout, was used to evacuate the oral cavity.

The levels of PM2.5 and PM10 particles were recorded prior to starting the procedure and every 10 second for 90 seconds. The time taken to return to the same level of PM2.5 and PM10 was also recorded. Particles meters were measured use using 3 identical commercially available particle meters; Huma-I Advanced Portable Air Quality Monitor (HI-150,) particle meter (Humatech, Bokhap-Munhwa Center, Incheon, Korea)

The 3 particle meters were located on a rig to record the airborne particles at the position of the clinician’s face, nurse’s face and patient’s chest (Figure 1). The clinician meter was 50cm above and 10cm behind the supine patient’s oral cavity. The nurse particle meter 50 above and 20cm to the left from the supine patient’s oral cavity. The patient chest meter was 10cm horizontal from the oral cavity in the long axis of the supine patient The meters were allowed to warm up and stabilize alongside each other in still air to ensure the base reading were identical before commencing each experimental sequence.

**Figure 1.**
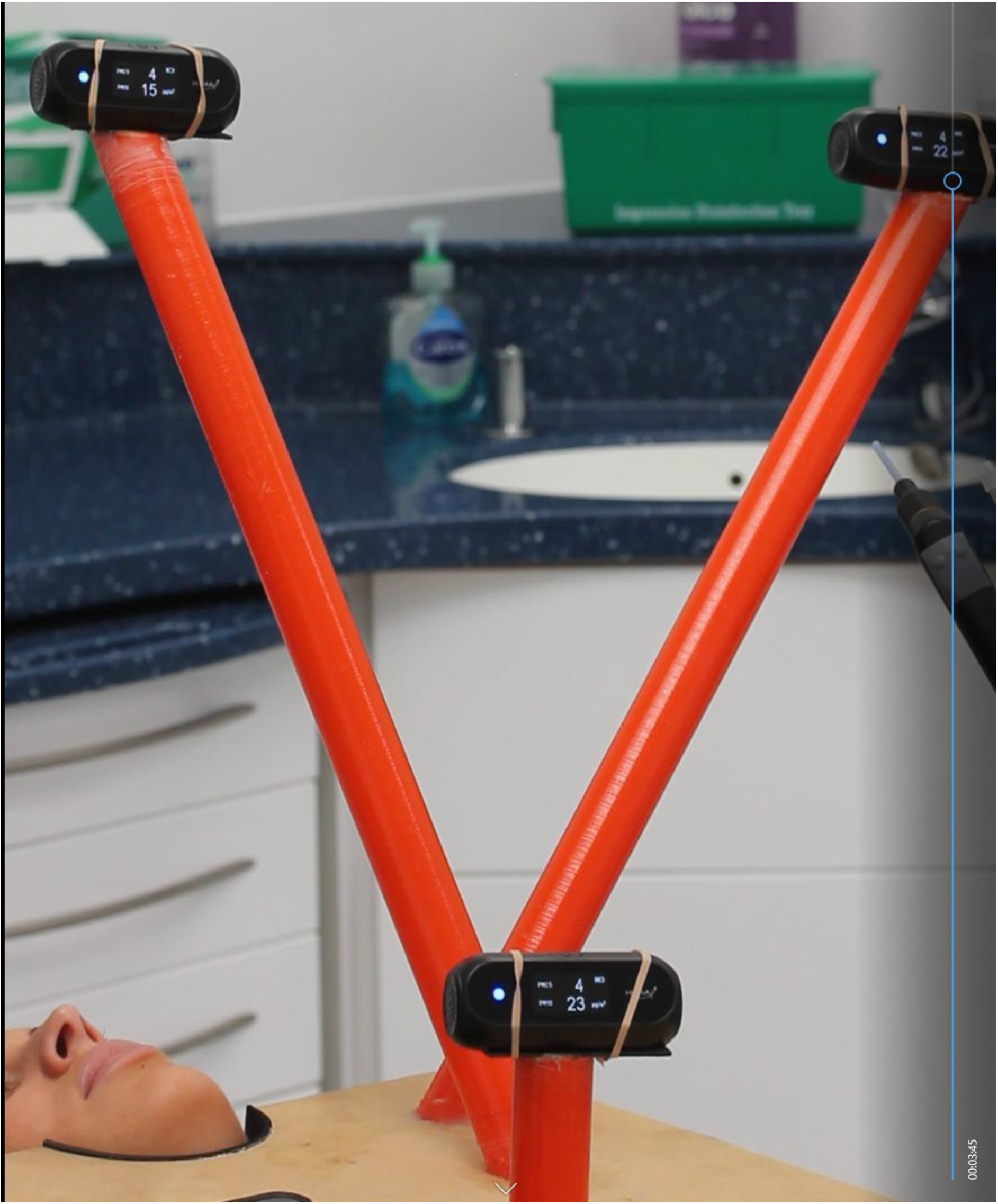
**Position of particle meters**

Three d-AGPs were undertaken as follows in the sequences:

1. 3-in-1 water/air syringe Operated into the oral cavity continuously for 90 seconds. Moving the spray tip clockwise around the dental arch, in both upper and lower arches
2. Air rotor. High speed air rota with a 1mm diameter diamond fissure bur, maximum water cooling for 90 seconds. Moving the bur clockwise around the dental arch, in both upper and lower arches next
3. Ultrasonic scaler Ultrasonic scaler, maximum water cooling for 90 seconds. Moving the scaler tip clockwise around the dental arch, in both upper and lower arches next

The experimental procedures were recorded by video so that all 3 meters were visible allowing replay and recording of data using the frame timeline. During all procedures, a 12mm internal diameter high volume suction tip was operated at maximum suction and held so the orifice was buccal to lower left molar. The water generating instrument (3-in-1, air rotor, ultrasonic scaler) was operated constantly with water flow at the maximum and moved around the upper and lower dental arch just above the incisal edge of the dentition, each circumnavigation taking about 10 seconds for a timed 90 second period.

Upon completion, both suction and water generating instrument were removed from the oral cavity and both patient and operator remained stationary until the particle meter reading for all 3 recording stations returned to original pre d-AGP base levels for at least 20 seconds for both PM2.5 and PM10. The time from stopping instrument use and returning to this level was recorded as the Time to Dissipate (TD) aerosol

The sequence of d-AGP procedures was repeated 3 times in each surgery, having allowed at least 5mins from when the PM2.5 and PM10 reading had returned to pre-d-AGP levels.

5 different surgeries were used in the data collection.

Practice 1 (surgery 1): 3.2×3.8×2.4m surgery with no ventilation either by extraction or window with door closed.

Practice 2 (surgeries 2 and 3): 3×4×2.4m surgeries with powered 4in extraction vent sited vertically above the patients head and open door behind operating clinical team which opening in to service corridor.

Practice 3 (surgery 4): 4.5×4×2.5m with closed door and no active ventilation. Surgery 5 - 4. x3.5×2.5m with closed door and no active ventilation.

The mean PM for 2.5 and 10 for all 3 situations was calculate and plotted as mean values with 95% confidence intervals for dentist, nurse and patient for each d-AGP and the mean time for PM2.5 and PM10 to return to pre-d-AGP levels.

## Results

As there was minimal difference in the readings for both PM2.5 and PM10 for all three sampling positions, the date was combined and presented as the mean value with upper and lower Confidence Intervals for PM2.5 and PM10 exposure in Figures 2-4.

**Figure 2.**
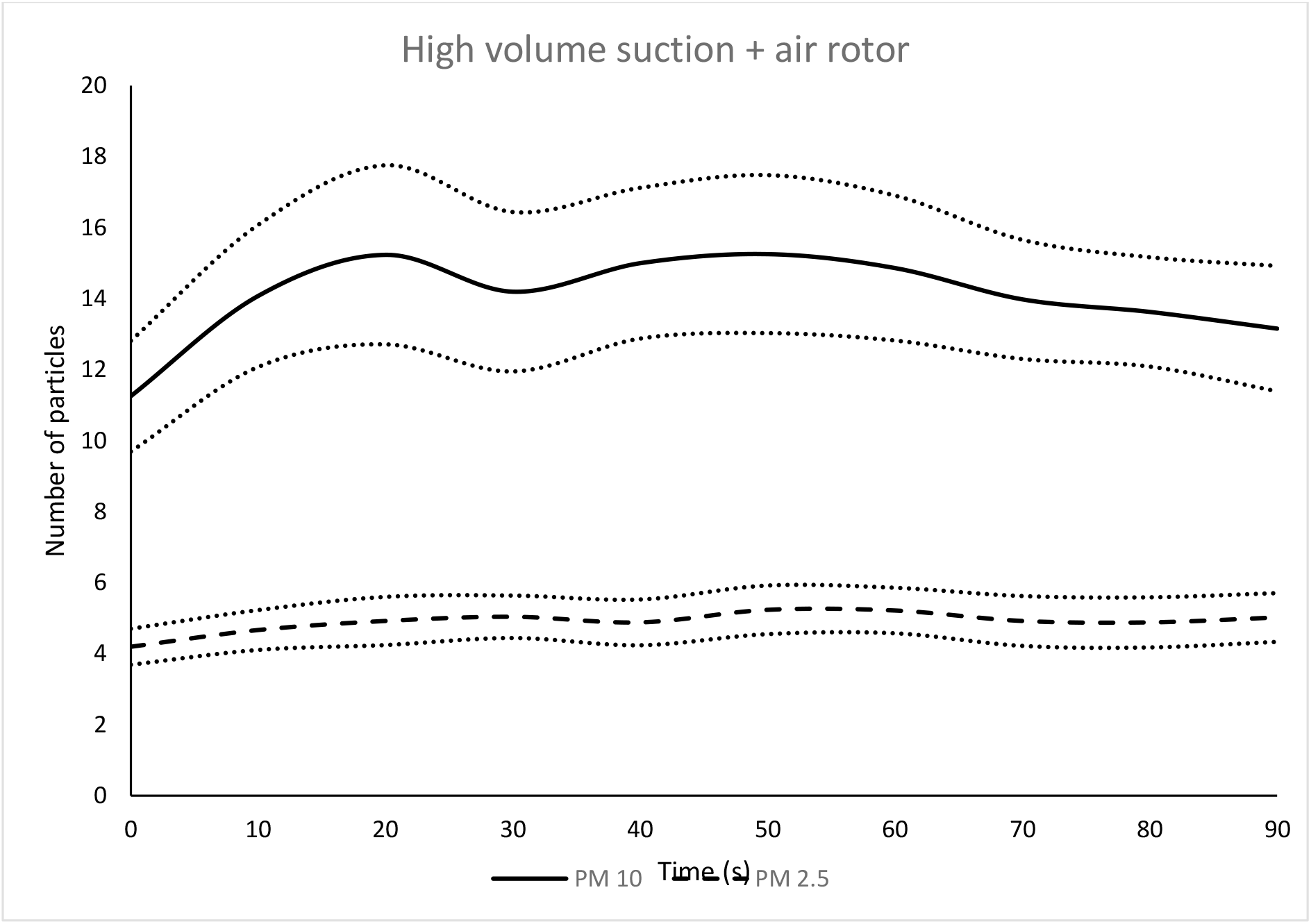
Air Rotor and High-Volume Suction: Mean number of particles recorded at 10 second intervals PM2.5 (dotted line) and PM10 (solid line) in _u_g/m^3^

**Figure 3.**
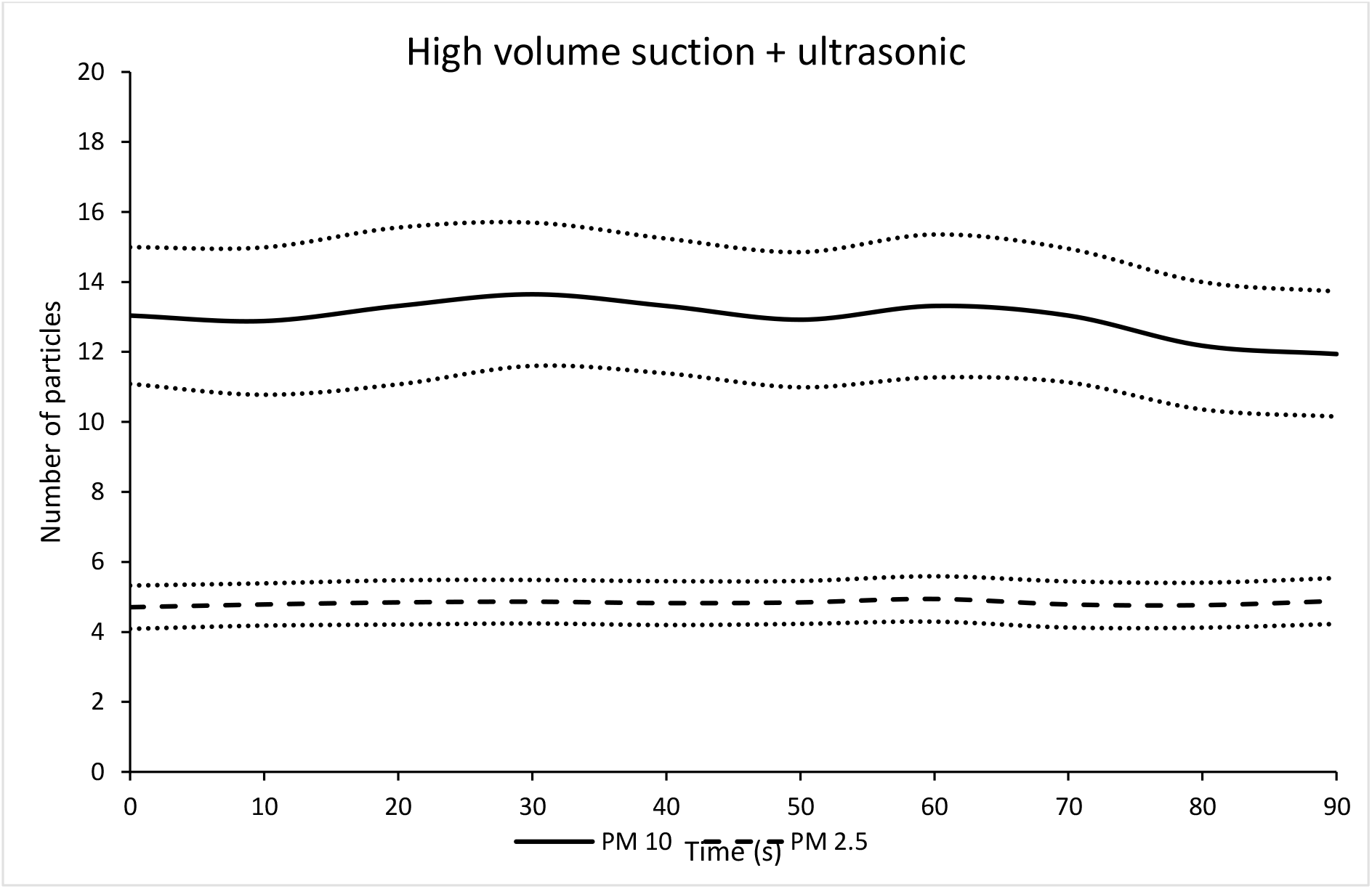
Ultrasonic scaler and High-Volume Suction: Mean number of particles recorded at 10 second intervals PM2.5 (dotted line) and PM10 (solid line) in in _u_g/m^3^

**Figure 4.**
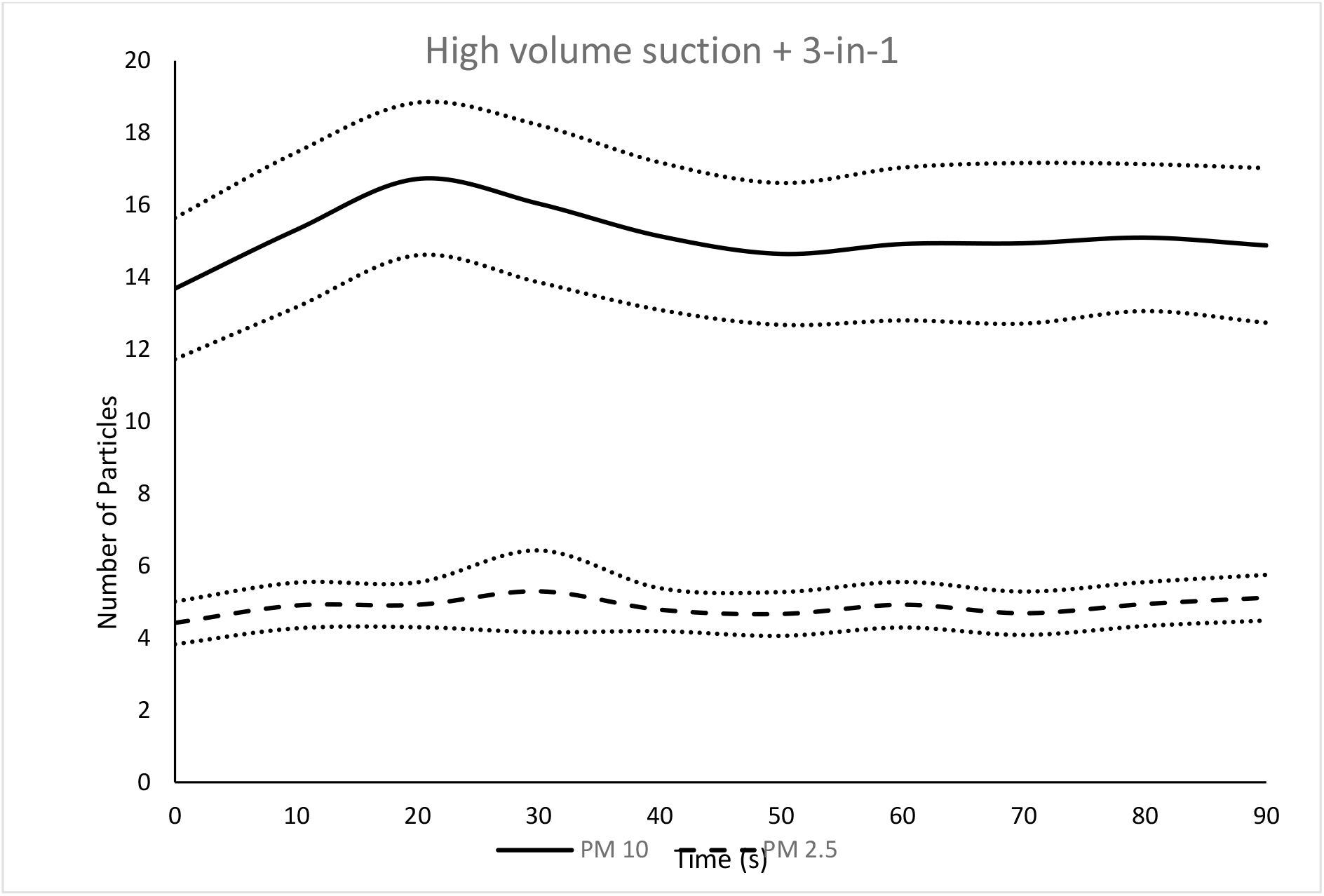
3-in-1 water/air syringe and High-Volume Suction: Mean number of particles recorded at 10 second intervals PM2.5 (dotted line) and PM10 (solid line) in in _u_g/m^3^

It can be seen that in all procedures there is an initial increase in PM10 particles, which stabilises within 30 seconds of starting the d-AGP. The increase is greatest with the use of the air rotor (Figure 2). It is interesting that the ultrasonic scaler (Figure 3) had the lowest increase in PM10. The increase is approximately 2-3 _u_g/m^3^ during the use of both air rotor and 3-in-1 water-air syringe and less than 1 _u_g/m^3^ during the use of the ultrasonic scaler In all procedures the PM2.5 measurement remain very near baseline throughout.

The time taken for PM2.5 and PM10 to return to the background reading was an average of 2 minutes 50 seconds (95% CI: 2:34 to 3:37).

## Discussion

This study was undertaken in primary care settings as it reflects the ‘real world’ that the vast majority of clinicians work in. The study limitations were the use of commercial, non calibrated particle meters. The meters were check alongside each other before each recording session to ensure reading were identical before commencing and the data was presented as the ‘change in readings’ as it is the change from background that is of interested when determining potential risk and length of fallow period required. Small aerosol particles are thought to have a greater role in airborne transmission of COVID19 infection due to being inhaled deeper into the respiratory tract, PM10 form part of the fomite transmission process. The particle meter reading for PM2.5 and PM10 are surrogates for the large and small aerosol particles generated during dental procedures. PM2.5 represent the smaller particles that are thought to penetrate more deeply into the respiratory system, PM10 are larger particles that are thought to coalesce and fall under gravity but can be inhaled into the upper respiratory tract.

The finding that PM2.5 levels remain extremely near background level indicated that dental AGPs when used with high volume suction release little, if any, additional small particles that potentially could be inhaled deep into the respiratory system. The increase of PM10 particles is also low

The increase large particles (2 to 3 μgm/m^3^) during the use of both the air rotor and 3-in1 water-air syringe is as expected, however the very low change in PM10 when using the ultrasonic scaler is a new finding and would suggest this presents a lower risk than previously thought. In all procedures the increase was very low being in the range of 2 to 3 μgm/m^3^ and this should be recognised.

The mean length of time for PM10 and PM2.5 to return to background levels was 2m 50s. This is greatly lower than that prosed by the UK Department of Health which recommend fallow periods of up to 60 minutes (OCDO, 2020, Clarkson et al., 2020). Around the world much shorter fallow periods are being used, if any; the majority of dental clinics worldwide using 10 minutes or less fallow period

These findings suggest that dental staff are at lower risk of catching COVID19 when undertaking dental AGPs than modelling proposes and this is supported by the empirical evidence from around the world where dental staff are at the same or lower level of risk of infection than the general population.

## Conclusions

Dentally generated aerosols are of very low levels, particularly of the smaller particles which are thought to be the possibly the main cause of airborne transmission of COVID19. This would suggest that dental staff are at a lower risk of COVID19 infection via dental AGPs than guidelines recommend and that a shorter fallow period of 10minute or less, is indicated.

## Data Availability

All data is available on request

## Notes

### Competing Interest Statement

The authors have declared no competing interest.

### Funding Statement

no funding

